# Selecting pharmacies for COVID-19 testing to ensure access

**DOI:** 10.1101/2020.09.17.20185090

**Authors:** Simon Risanger, Bismark Singh, David Morton, Lauren Ancel Meyers

**Affiliations:** Department of Industrial Economics and Technology Management, Norwegian University of Science and Technology, Trondheim, Norway; Department of Mathematics, Friedrich-Alexander-Universität Erlangen-Nürnberg, Erlangen, Germany; Department of Industrial Engineering and Management Sciences, Northwestern University, Evanston, IL, USA; Department of Integrative Biology and Department of Statistics and Data Sciences, The University of Texas at Austin, Austin, TX, USA

## Abstract

Rapid diagnostic testing for COVID-19 is key to guiding social distancing orders and containing emerging disease clusters by contact tracing and isolation. However, communities throughout the US do not yet have adequate access to tests. Pharmacies are already engaged in testing, but there is capacity to greatly increase coverage. Using a facility location optimization model and willingness-to-travel estimates from US National Household Travel Survey data, we find that if COVID-19 testing became available in all US pharmacies, an estimated 94% of the US population would be willing to travel to obtain a test, if warranted. Whereas the largest chain provides high coverage in densely populated states, like Massachusetts, Rhode Island, New Jersey, and Connecticut, independent pharmacies would be required for sufficient coverage in Montana, South Dakota, and Wyoming. If only 1,000 pharmacies in the US are selected to provide testing, judicious selection, using our optimization model, provides estimated access to 29 million more people than selecting pharmacies simply based on population density. COVID-19 testing through pharmacies can improve access across the US. Even if only few pharmacies offer testing, judicious selection of specific sites can simplify logistics and improve access.

## 1 Introduction

Mass testing and contact tracing for COVID-19 will be critical to mitigate the impact of the disease prior to the availability of a vaccine [17, 30]. While South Korea and Germany quickly launched expansive testing programs [5, 22], efforts are rolling out more slowly in the UK, Italy, and the US. In Guidelines for Opening Up America Again, the White House and the Centers for Disease Control and Prevention (CDC) provide benchmarks for states prior to relaxing COVID-19 social distancing orders [23]. A key criterion is “the ability to quickly set up safe and efficient screening and testing sites for symptomatic individuals and trace contacts of COVID+ results” [23]. Most states are expanding COVID-19 testing capacity at a diverse array of test sites, including commercial labs, healthcare clinics, hospitals, pharmacies, schools, churches, and sports complexes [11, 13, 15]. Although the US has increased its testing activity, fair access to testing remains a challenge. Data from New York City shows that people in poor and immigrant neighborhoods were less likely to be tested, despite having larger likelihood of testing positive [2]. According to the CDC, current data suggest that racial and ethnic minority groups have a disproportional burden of illness [4].

There are two classes of SARS-CoV-2 tests: viral tests that assay for current infection and antibody tests that detect prior infection. Viral testing allows for contact tracing and case isolation strategies to contain emerging clusters of cases [12]. Individuals infected with SARS-CoV-2 are thought to be infectious several days before they feel symptoms [7]. Thus, wide testing of all who believe they may be at risk for infection can improve pandemic mitigation [16]. However, when testing capacity is limited, individuals with COVID-like symptoms or known exposure are prioritized [9, 21].

Here, we argue that a state or federally coordinated effort to offer testing through pharmacies could rapidly address gaps in coverage in the US. Pharmacies, which are ubiquitously found in both drug stores and grocery stores, are well positioned to expand access to COVID-19 testing. First, many people feel more comfortable walking into a pharmacy than a local hospital or outpatient clinic, and second, many live closer to a pharmacy than a healthcare facility [3, 14]. Over 90% of Americans live within five miles of a pharmacy [3]. Third, pharmacies have a skilled workforce that can be utilized to administer testing [22]. Some pharmacies also have walk-in clinics. As of 2017, there were an estimated 2,800 retail clinics in the US, with three quarters operated by CVS or Walgreens [18]. Fourth, large pharmacy chains have extensive supply-chain networks and logistics infrastructure, with stores located throughout a state. These distribution networks were leveraged during the 2009 H1N1 pandemic by several states to distribute federally allocated antivirals [1]. Finally, the Association of State and Territorial Health Officials (ASTHO) recognizes pharmacies as instrumental for immunization campaigns and administering vaccines [1]. Given the vital role pharmacies could play in rapidly expanding COVID-19 testing capacity and accessibility, the U.S. Department of Health & Human Services (HHS) approved pharmacies to order and administer tests for COVID-19 as of April 8, 2020 [26].

Pharmacies are already engaged in testing. HHS has established a public-private partnership with selected pharmacies and retailers. Their goal is to provide more accessible and convenient testing and to ensure protection of healthcare personnel [27]. As of July 2020, selected locations from CVS, Kroger, Rite Aid, Walgreens, Walmart, and independent pharmacies in partnership with Health Mart and eTrueNorth, are offering SARS-CoV-2 tests [27]. Pharmacies are also scaling up their own testing beyond the HHS-sponsored community-based testing sites. CVS and Walgreens offer drive-through testing in several states, with the intention of expanding testing locations [6, 29].

The expansion of pharmacy-based COVID-19 testing would not only help to bridge existing gaps between current capacity and national reopening targets, but also provide access for hard-to-reach and at-risk communities. Here, we estimate the geographic coverage that could be achieved through a pharmacy-based testing program, in terms of the proportion of individuals who would be willing to travel to the nearest pharmacy testing site to obtain a COVID-19 test (if warranted), and the distance they would be required to travel. We use an optimization model to determine specific pharmacy locations in a given state or nationwide that would provide the greatest access. Our findings indicate whether and where major chains can provide sufficient access on their own.

## 2 Methods

We formulate and solve an optimization model to select locations for pharmacies. The model is similar to that of Singh et al. [20], who used it to support the distribution of strategic national stockpile antivirals during pandemics. The model requires as input a geographic region – e.g., a US state – partitioned into ZIP code areas. Given a specific number of locations (ZIP code areas) to be selected, solving the optimization model yields the subset of store locations that maximizes the expected number of people willing to travel to their closest pharmacy, among the selected locations. That is, the optimization model maximizes coverage of the population given a limited “budget” of locations, which can be selected. We solve the optimization model parametrically in this budget to understand the trade-off between the total number of locations and coverage.

Mathematically, we represent the problem as the facility location optimization model expressed in formulation (1). Table 1 outlines the nomenclature. The objective (1a) is to maximize the population willing to travel to its closest pharmacy. Index *i* ∈ *I* denotes all areas (ZIP codes), while *j* ∈ *J* ⊆ *I* denotes areas with pharmacies. The population willing to travel from area *i* to its closest pharmacy is the product of the population of area *i*, *P_i_*, the fraction of population willing to travel between areas *i* and *j*, *ρ_ij_*, and variable *y_ij_*, which declares the area *j* closest to *i*. Constraint (1b) sets the budget of locations, where *N* is a hyperparameter and variable *x_j_* expresses whether a pharmacy in area *j* is selected or not. Constraint (1c) finds the pharmacy closest to area *i*. The model can only select one *y_ij_* for each area *i*, and consequently chooses the *y_ij_* that maximizes objective (1a). Constraint (1d) ensures that the closest pharmacy is open, while (1e) and (1f) force *x_j_* and *y_ij_* to be binary.

**Table 1:** Nomenclature for optimization problem (1).

| **Sets and indices** |  |
| --- | --- |
| $i \in I$ | Set of all areas |
| $j \in J \subseteq I$ | Set of areas that contain a pharmacy |
| **Parameters** |  |
| $P_i$ | Population of area $i$ |
| $\rho_{ij}$ | Fraction of population willing to travel from area $i$ to area $j$ |
| $N$ | Maximum number of pharmacy areas |
| **Decision variables** |  |
| $x_j$ | Select pharmacy of area $j$ or not |
| $y_{ij}$ | Closest pharmacy area $j$ for area $i$ |

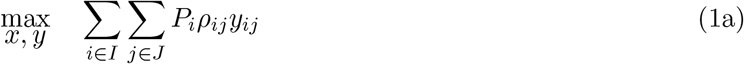

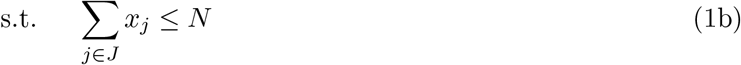

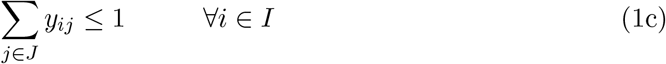

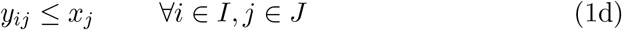

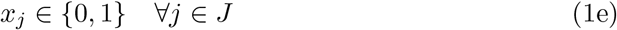

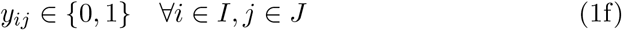

The model require the willingness to travel of the population as input. We use the data-driven willingness-to-travel function from [20], which is estimated using National Household Travel Survey data [28] for privately operated vehicles. Equation (2) shows the fraction of the population, *ρ*, willing to travel distance *d*. The willingness-to-travel function is an exponentially decaying fraction of the population, where the population has different travel behavior for distances below and above a five-mile threshold. This threshold considers different mobility patterns in urban and rural areas, where the willingness to travel has a steeper decrease below five miles. We calculate parameter *ρ_ij_* as *ρ*(*d_ij_*) where *d_ij_* is the distance between area *i* and *j*.

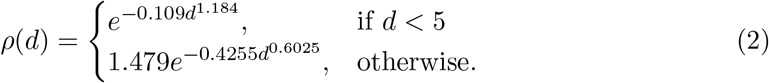

For areas, we use a mapping from ZIP codes to ZIP Code Tabulation Areas (ZCTAs). The former represents a set of mail delivery routes, while ZCTAs correspond to a geographic area used by the US Census Bureau. Population counts, centroid coordinates, and land areas for each ZCTA are obtained from the 2010 US Census. The US Census provides coordinates, latitudes and longitudes, for the centroid of each ZCTA. We calculate travel distance *d_ij_* as the distance between two ZCTA centroid coordinates, as calculated by the Haversine formula. As in Singh et al. [20], we assume that the full population in a ZCTA is willing to travel to a pharmacy in their own ZCTA. The travel distance to pharmacies within a ZCTA is approximated as the square root of land area of the ZCTA divided by two for the full population. For pharmacies, we use a national dataset for operating pharmacies in the US, produced by InfoGroup and Amazon Web Services in coordination with Esri’s national COVID-19 response team.

Note that the optimization model only considers whether a pharmacy is present in a particular ZCTA. Hence, the model does not consider the capacity of each pharmacy to provide diagnostic testing. This reduces data requirements to run the model, and helps improve its computational tractability. In effect, we assume a pharmacy location at the centroid of each ZCTA. Formulation (1) requires a decision variable *y_ij_* between each ZCTA and pharmacy location. This becomes computationally burdensome for large states and national model instances. We therefore only include variables for combinations that have at least a 20% willingness to travel. A post-processing step connects ZCTAs with less than a 20% willingness to travel to their closest pharmacy. We implement formulation (1) in Julia, using the open source package JuMP [8] and the Gurobi solver.

## 3 Results

Our analysis considers 67,473 pharmacies within the US. CVS, Walgreens, and Walmart are the three largest chains, and operate 15.1%, 13.0%, and 6.9% of these stores, respectively. We consider two scenarios: a nation-wide program administered federally and state-wide programs administered separately by the 50 states.

We estimate that a national pharmacy-based testing program using the entire network of major chains and independent stores would provide access for 94.0% of the US population. Section A in the online supplement provides detailed results. Collectively, all pharmacies, both chains and independents, operate stores in 14,474 unique ZCTAs nationwide, which is less than half of the 32,930 ZCTAs in the US Census. The resulting median distance from home to the nearest pharmacy-based COVID-19 testing site would be 2.4 miles, with 90% of the population living within 7.4 miles of the nearest testing site.

Reducing the number of pharmacy test sites would reduce access, but not by much if the pharmacy locations were specifically selected to ensure broad access. For example, 10,000 geographically dispersed stores – chosen via the optimization model to maximize willingness to travel – would be expected to provide access to 90.3% of the US population, with a 2.5-mile median travel distance, and 90% of people living within 8.0 miles of the nearest COVID-19 testing site. Half as many pharmacy test sites (located in 5,000 ZCTAs) would reduce expected access to 77.7% while increasing the median and the 90th percentile travel distance to 3.1 miles and 10.8 miles, respectively.

To simplify coordination and logistics, a policymaker could rely on a single major chain. Although none of the major chains can reach over 80% of the US population, Table 2 shows that the median travel distance to the nearest test site would be expected to remain under five miles. Any pair of the major chains – CVS, Walgreens, and Walmart – could provide 80% access. CVS and Walgreens could reach an estimated 84.2% of the population; adding Walmart would slightly expand coverage to 86.5%, but still fall short of the 94.0% estimated if all US pharmacies offer testing.

**Table 2:** Estimated proportion of the US population willing to travel to the nearest pharmacy and corresponding travel distances

| Chain | Population access | Travel distance [miles] |  |  |
| --- | --- | --- | --- | --- |
|  |  | 10th percentile | Median | 90th percentile |
| All | 94.0% | 1.0 | 2.4 | 7.4 |
| All (max 10,000 locations) | 90.3% | 1.1 | 2.5 | 8.0 |
| All (max 5,000 locations) | 77.7% | 1.4 | 3.1 | 10.8 |
| CVS | 75.5% | 1.1 | 2.9 | 15.4 |
| Walgreens | 76.4% | 1.2 | 3.0 | 13.5 |
| Walmart | 65.2% | 1.8 | 4.5 | 12.1 |
| CVS & Walgreens | 84.2% | 1.0 | 2.6 | 10.4 |
| CVS & Walmart | 82.6% | 1.1 | 2.8 | 10.1 |
| Walgreens & Walmart | 81.8% | 1.2 | 2.8 | 10.2 |
| CVS, Walgreens & Walmart | 86.5% | 1.0 | 2.6 | 9.3 |

We also analyzed the coverage of state-level pharmacy testing sites. See Section B in the online supplement for details. If all chain and independent pharmacies provide COVID-19 testing, we estimate that 47 states would achieve at least 80% access. Only Vermont (77.3%), Alaska (78.0%), and Maine (79.9%) fall short. Access is estimated to exceed 95% in 12 states, including the most populous states of California (97.8%), Florida (97.3%), New York (95.8%), and Texas (95.0%).

If states were to offer testing through just a single chain, we estimate that CVS, Walgreens, and Walmart stores could reach over 80% of the population in 17, 13 and seven US states, respectively. Single chains can provide far greater access to COVID-19 testing in densely populated states than in sparsely populated states. For example, in Massachusetts, Rhode Island, New Jersey, and Connecticut, CVS – the largest pharmacy chain in these states – achieves over 80% access, with half of the population traveling no more than 2.3 miles to their nearest CVS store. In contrast, the largest pharmacy chain in Montana, South Dakota, and Wyoming would be expected to provide accessible testing to under 50% of the respective populations, with half living over 9.0 miles and 10% living over 50 miles from the nearest store in the chain.

As of July 2020, there is not a comprehensive listing of US testing sites. However, Georgia, Illinois, and Minnesota provide state-level lists [11, 13, 15]. Table 3 outlines the results. With locations per 28 July 2020, we estimate that testing locations in Illinois and Minnesota are accessible to slightly above 70% of the state populations, while Georgia’s site cover just over 50% of the population. If testing were offered at all chain and independent pharmacies, all three states can significantly increase access and reduce travel distance. The largest pharmacy chain in Georgia and Illinois would be expected to provide better coverage than current sites, especially in Georgia where CVS stores could provide access to over 80% of the population. All three states could significantly increase their access by adding the the locations of the largest pharmacy chain to their current testing sites.

**Table 3:** Estimated access in three states from existing test sites (per 28 July 2020) [11, 13, 15] and pharmacies (all and largest chain) with travel distances.

| State | Distribution | Population | Travel distance [miles] |  |  |
| --- | --- | --- | --- | --- | --- |
|  |  | access | 10th percentile | Median | 90th percentile |
| Georgia | All pharmacies | 94.6% | 1.8 | 3.4 | 7.4 |
|  | Existing test sites | 53.5% | 2.8 | 6.5 | 13.3 |
|  | CVS | 80.5% | 1.8 | 3.7 | 10.7 |
|  | CVS & existing | 86.1% | 1.7 | 3.6 | 8.4 |
| Illinois | All pharmacies | 94.9% | 0.9 | 1.8 | 5.8 |
|  | Existing test sites | 73.5% | 1.0 | 2.6 | 10.7 |
|  | Walgreens | 86.8% | 0.9 | 1.9 | 9.1 |
|  | Walgreens & existing | 88.8% | 0.9 | 1.8 | 7.4 |
| Minnesota | All pharmacies | 89.5% | 1.1 | 3.4 | 9.0 |
|  | Existing test sites | 73.7% | 1.3 | 4.4 | 12.3 |
|  | Walgreens | 70.8% | 1.2 | 3.9 | 23.5 |
|  | Walgreens & existing | 81.8% | 1.2 | 3.9 | 11.3 |

## 4 Discussion

As US states ramp up COVID-19 testing to allow broad and fair access, our results suggest that pharmacy-based testing would provide good coverage and reasonable travel distances for large portions of the US population. We provide an actionable strategy for selecting specific pharmacy test sites to ensure access, given that the density and proximity of pharmacies varies enormously across the US. We have applied the strategy to demonstrate the potential reach of national-level and state-level pharmacy testing programs. Our approach can likewise be used to select pharmacies on a local scale or to close known gaps in coverage by ongoing testing efforts.

During the 2009 H1N1 influenza pandemic, pharmacies worked with state health departments to distribute strategic national stockpile antivirals [1]. Our findings suggest that tailoring testing strategies to individual states could achieve broader coverage than a onesize-fits-all national strategy. Densely populated states, like Rhode Island, Massachusetts, and New York may be able simplify testing logistics by coordinating only with a single chain. In less densely populated states, like Montana, South Dakota, and Wyoming, enlisting more than a single pharmacy chain would be required to reach more than half the population.

For the majority of states that fall between these two extremes, judicious selection of pharmacy locations is important but not obvious. Our optimization approach would help such states to provide the broadest possible access to COVID-19 testing while keeping the coordination simple. Consider, for instance, Illinois and Texas. Both have densely populated metropolitan areas and significant rural populations. Our analysis suggests a viable strategy that would work well for both. First, identify the major chain with the best geographic reach – Walgreens provides an estimated 86.8% coverage in Illinois and CVS provides an estimated 80.6% coverage in Texas – and then selectively target single independent pharmacies to cover the remainder of the largely rural population. An additional 50 independent pharmacies in Illinois and 182 in Texas would be expected to raise coverage to 90%.

Our approach is simple and effective. As a benchmark, consider an alternative in which pharmacies are selected as testing sites based simply on local population density without considering travel distances. If we select testing sites in only 1,000 ZCTAs, then this heuristic would choose pharmacies primarily in dense urban areas and thereby provide access to an estimated 39.3% of the US population. In contrast, our approach identifies pharmacies that would be expected to increase coverage to 48.1%, and hence provide access to an estimated additional 29 million people. For 5,000 pharmacies, our approach again beats this heuristic, raising expected coverage from 73.9% to 77.7% of the US population, a difference of 12.5 million people. The benefits of our pharmacy selection strategy are amplified when resources are scarce and coverage is limited.

We estimate that, as of 28 July 2020, COVID-19 test sites in Georgia barely provide access for half of the state population and test sites in Illinois and Minnesota miss over a quarter of their populations. Georgia reports 150 test sites, and if they were to provision COVID-19 tests through all 2,345 pharmacies in the state (or alternatively, one in each of the 424 ZCTAs that have a pharmacy), estimated access would increase from 53.5% to 94.6% of the population. Offering testing through the single largest pharmacy chain in the state (CVS) in addition to current sites would increase estimated coverage to 86.1%. Ramping up testing in Walgreens stores in Illinois and Minnesota would also narrow the gap in coverage, but to a lesser extent.

We make a critical assumption that may bias our estimates of people’s willingness to travel to nearby pharmacies for COVID-19 test when symptoms present. Using data from the National Household Travel Survey [28], we assume that individual willingness to travel for diagnostic testing coincides with willingness to travel by privately operated vehicle for work, school, family or social reasons under normal, non-emergency, conditions. Our estimates may be pessimistic, given the urgency of COVID-19, or possibly optimistic, if COVID-19 symptoms reduce willingness to leave the home. Still, surveys suggest that the US population is willing to seek COVID-19 tests. In one such survey, 70% claim that they are willing to take a free test [24], while 80% are willing to pay less than $2 for at-home antibody test kits [19]. If our estimates are incorrect but in a consistent manner across the US population – for example, if willingness to travel is higher than we estimate by a consistent proportion – then the estimated coverage will be similarly incorrect, but the optimal selection of pharmacies would remain the same.

We also note that our analysis considers the location (ZCTA) but not the capacity of each pharmacy. As outlined in Section 2, the optimization problem does not distinguish single versus multiple pharmacies in a ZCTA. Commercial pharmacy capacity may be sized to handle local demand under normal operations but may not suffice to handle a surge in demand. Nevertheless, policymakers can open additional pharmacies or other sites in high demand areas if they experience insufficient capacity.

Our pharmacy selection methodology can flexibly accommodate plausible policy constraints. For example, we can identify specific stores within chains that have already launched testing programs to expand access to under-served populations. If states have established targets for accessibility of COVID-19 testing, we can determine the minimum number and best locations of pharmacies required to achieve such targets. When recommending additional pharmacy testing sites, the method can account for existing COVID-19 testing capacity through pharmacies, labs, healthcare systems, public health centers and other sites. It can also exclude chains or individual stores from consideration, such as pharmacies co-located with grocery stores to reduce exposure to healthy shoppers, or pharmacies without adequate facilities or staff.

Willingness-to-travel estimates can provide insight into geographic and socioeconomic disparities in access to diagnostic tests and other pharmacy-provided resources during the COVID-19 pandemic. For instance, Fouad et al. [10] show that COVID-19 affects African Americans disproportionately and recommends ensuring equitable testing, while Tsai and Wilson [25] explain how COVID-19 can be a public health problem for homeless populations. Our approach can rapidly identify candidate pharmacy-based testing sites to ameliorate such gaps.

## 5 Conclusion

Although testing capacity has been increasing since February 2020, many communities still lack adequate access to COVID-19 tests in the US. The bottleneck is not just the quantity of tests, and the ability to process tests, but also the number, staffing and locations of test sites. To meet these needs, pharmacies have selectively begun to ramp up COVID-19 testing. Our results suggest that broad and coordinated pharmacy-based testing programs can provide estimated access to 94% of the US population. Even a subset of pharmacies, for instance pharmacies of a specific chain, could ameliorate critical gaps in coverage. The facility location optimization is a simple method for selecting locations to improve access, which can adjust to policy constraints. For instance, densely populated states can provide acceptable access through a single pharmacy chain, while other states can use the optimization framework to judiciously select locations to maximize cover. As America continues to ‘open up’, robust and expansive testing, contact tracing, and isolation programs will be key to mitigating future transmission and averting unmanageable surges in COVID-19 hospitalizations. Our procedure for determining testing sites can be easily applied to fill in persistent geographic and socioeconomic gaps in coverage across the US.

## Data Availability

ZIP Code Tabulation Areas and corresponding 2010 population counts, centroid coordinates,
and land areas are openly available by the US Census Bureau. National Household Travel Survey data used to estimated willingness to travel is openly available from the US Department of Transportation. The national dataset for operating pharmacies in the US was produced by InfoGroup and Amazon Web Services in coordination with Esri's national COVID-19 response team and shared with us for this study.

## Acknowledgements

The authors thank Dr. Gordon Wells for acquiring and sharing the pharmacy data set used in this study, and for providing additional pharmacy data sets that we considered. The authors are also grateful to John Sheffield and Mauricio Tec, who helped gather and share pharmacy data sets. We acknowledge financial support from the National Institutes of Health under Grant NIH R01 AI151176 and Grant NIH U01 GM087791, the U.S. Department of Homeland Security under Grant 2017-ST-061-QA0001 and Tito’s Handmade Vodka. The views and conclusions contained in this document are those of the authors and should not be interpreted as necessarily representing the official policies, either expressed or implied, of the U.S. Department of Homeland Security.

